# Potential impact of individual exposure histories to endemic human coronaviruses on age-dependence in severity of COVID-19

**DOI:** 10.1101/2020.07.23.20154369

**Authors:** Francesco Pinotti, Paul S Wikramaratna, Uri Obolski, Robert S Paton, Daniel S C Damineli, Luiz C J Alcantara, Marta Giovanetti, Sunetra Gupta, José Lourenço

## Abstract

Cross-reactivity to SARS-CoV-2 from previous exposure to endemic coronaviruses (eHCoV) is gaining increasing attention as a possible driver of both protection against infection and severity of COVID-19 disease. Here, we use a stochastic individual-based model to show that heterogeneities in individual exposure histories to endemic coronaviruses are able to explain observed age patterns of hospitalisation due to COVID-19 in EU/EEA countries and the UK, provided there is (i) a decrease in cross-protection to SARS-CoV-2 with the number of eHCoV exposures and (ii) an increase in potential disease severity with number of eHCoV exposures or as a result of immune senescence. We also show that variation in health care capacity and testing efforts is compatible with country-specific differences in hospitalisation rates. Our findings call for further research on the role of cross-reactivity to endemic coronaviruses and highlight potential challenges arising from heterogeneous health care capacity and testing.

## Introduction

COVID-19 and its causative agent, SARS-CoV-2, have recently emerged as a global threat to human health, forcing many countries to undertake unprecedented measures to contain its spread. This disease displays a spectrum of illness severity and fatality characterised by a marked age gradient. Typically, cases under 20 years of age display mostly mild or no symptoms, while older individuals are at increased risk of developing severe symptoms, including respiratory failure, multiorgan dysfunction and death (1,2).

Understanding the determinants of severe symptoms is key to preparedness against COVID-19. So far, cohort studies have identified a number of risk factors for severe illness in comorbidities such as cardiovascular disease, diabetes mellitus and obesity (3−5). Meanwhile, there have been extensive efforts to calculate age-specific odds of developing clinical and severe symptoms, as well as hospitalisation and fatality rates (6−8). These results have important implications for public health, influencing real-time management and strategic allocation of clinical resources. Nonetheless, apart from a few notable exceptions (9−11), most modelling work assumes that SARS-CoV-2 spreads across an entirely susceptible population. Consequently, the impact of cross-reactivity between SARS-CoV-2 and other endemic human coronaviruses (eHCoVs), remains largely unexplored.

SARS-CoV-2 is the seventh coronavirus known to infect humans. SARS-CoV and MERS-CoV emerged in the past 20 years, while HCoV-229E, -NL63, -OC43 and -HKU1 are endemic to the human population. Infection with eHCoVs is frequent but, contrary to emergent HCoVs, it is usually associated with mild respiratory illness (12). Typically, the first exposure to any eHCoV occurs early during childhood, but reinfection can occur (13) due to the waning of homotypic immunity (14−17).

So far, a fully mechanistic explanation of COVID-19 severity, that accounts also for the heterogeneous immune landscape in which SARS-CoV-2 spreads, is lacking. T cell and IgG antibody reactivity to SARS-CoV-2 has been observed in non-exposed individuals (18−25), indicating that there is cross-reactivity between eHCoVs and SARS-CoV-2. It is still unclear, however, whether pre-existing cellular and antibody responses to SARS-CoV-2 are protective or pathogenic (26−32). In this study, we present a parsimonious model of eHCoV co-circulation to explore the effect of these contrasting possibilities on the age distribution of COVID-19 severity. The key assumption is that distinct life-histories of exposure to eHCoVs result in responses of varying effectiveness upon challenge by SARS-CoV-2 and, eventually, distinct clinical outcomes. We assume that the first infection with any eHCoV induces cross-protection against disease, but this is reduced in subsequent infections which more readily induce strain-specific responses. Furthermore, the risk of severe disease may increase with exposure to eHCoVs due to the selective amplification through immune priming of harmful responses such as antibody- and/or T-cell-mediated over-production of pro-inflammatory cytokines (33−36). We contrast the results of this model with one where risk of disease is exposure-independent (i.e. determined solely by factors such as immune senescence) and outline the conditions under which they provide a good fit to the age-specific hospitalisation rates in EU and European Economic Area (EEA) countries and the UK.

## Results

### eHCoVs calibration and dynamics

We represent the dynamics of the system using a multi-strain epidemic model with *5* coronavirus strains. We capture the immunological landscape prior to SARS-CoV-2 emergence by calibrating the model to available epidemiological knowledge (Figure 1) regarding the *4* known eHCoVs, HCoV-229E, -NL63, -OC43 and -HKU1; a fifth strain playing the role of SARS-CoV-2 is introduced only at a later stage (see Methods for more details).

**Figure 1:**
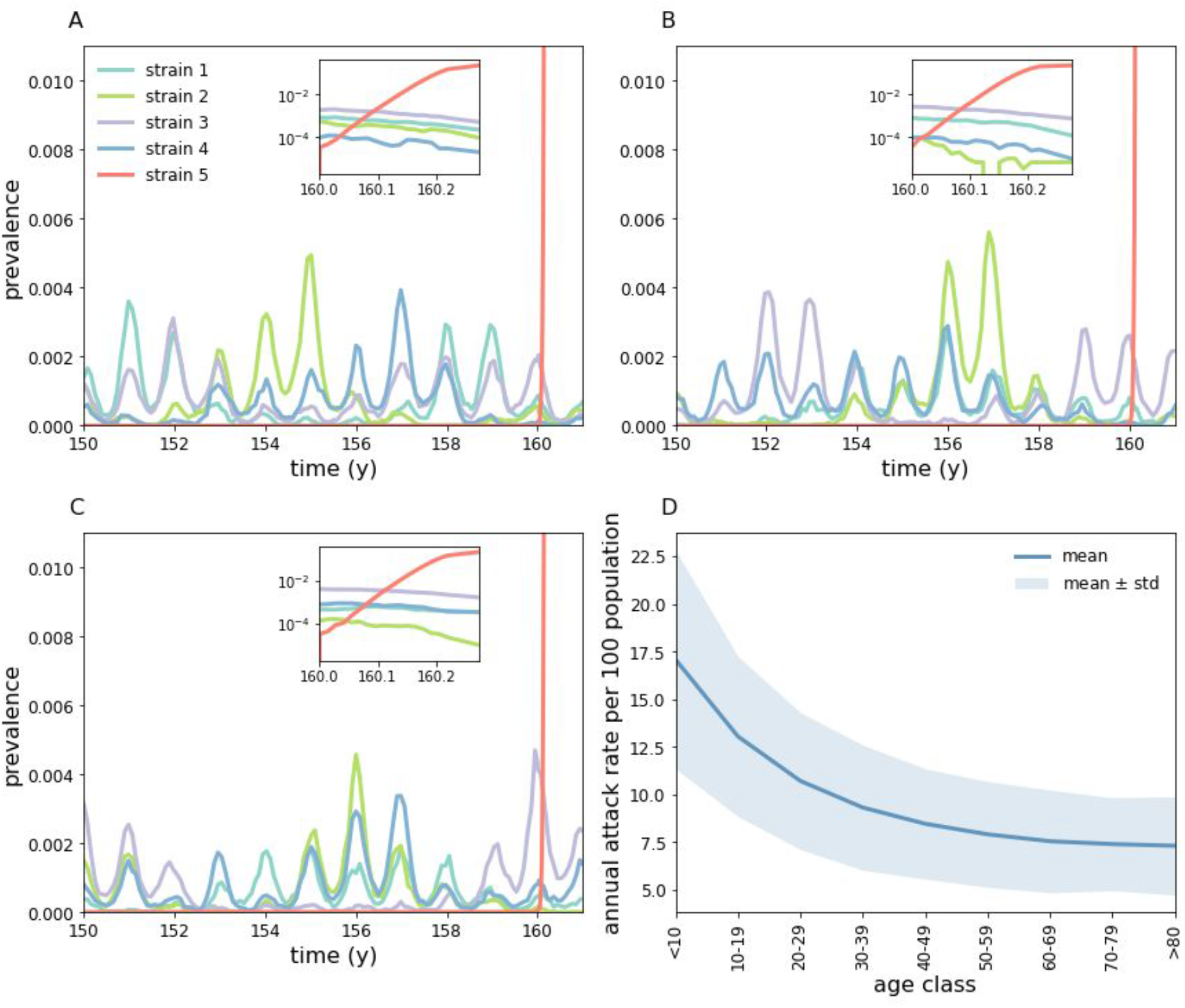
Simulated transmission of eHCoVs. (A-C) Stochastic realisations of the multi-strain model obtained using baseline parameters. At time *t* = *T*_*inv*_ = 160 *y* we randomly infect 10 individuals with the emergent strain (red). Insets: zoom on the first 100 days after introduction of the emergent strain. (D) Annual attack rate per 100 population by age class for all four eHCoVs. The attack rate decreases and plateaus with age since individuals accumulate immunity through consecutive infections with different eHCoVs.

Epidemiological parameters characterizing eHCoVs and SARS-CoV-2 were informed, where possible, from the literature. As reinfection is commonly observed in eHCoVs (15,37,38), we assume that the recovery provides only partial protection against reinfection with the same strain; in particular, we assume that exposure to a previously encountered strain results in infection with probability ρ. The case ρ = 0 corresponds to complete, life-long immunity upon recovery. We investigated the impact of the remaining parameters numerically, leveraging available epidemiological knowledge about eHCoVs to guide our analysis. Values defining the life-expectancy distribution were set to simulated age profiles matching average European patterns (simulated age profiles are shown in Figure S1). Model parameters and their values are briefly summarised in Table S1, while details about model implementation and calibration are found in Supplementary Note S1.

Figure 1A-C shows typical realizations of our multi-strain model under baseline conditions (parameter values indicated in bold in Table S1). The model captures annual patterns of eHCoV spread (39), with seasonal differences in eHCoV seasonal epidemics dictated mostly by stochasticity, population turnover and seasonal variation in transmissibility. At time *T* _*inv*_ = 160 *y* we introduce the emergent strain into the host population. As shown in the insets in Figure 1, the invading strain rapidly spreads through the population thanks to its antigenic novelty, infecting a large proportion of hosts.

We set susceptibility to reinfection to ρ = 0.35 in order for our model to yield a realistic force of infection (FOI) for eHCoVs. That is, we calibrated p in order to match empirical estimates of the age at first eHCoV infection (Figure 2), which is given by the inverse of the FOI (40). In our model, the emergent strain shares the same value of p as that of endemic strains. This assumption is compatible with recent work finding similar kinetics of antibody responses after both SARS-CoV-2 and eHCoVs infections (41).

**Figure 2:**
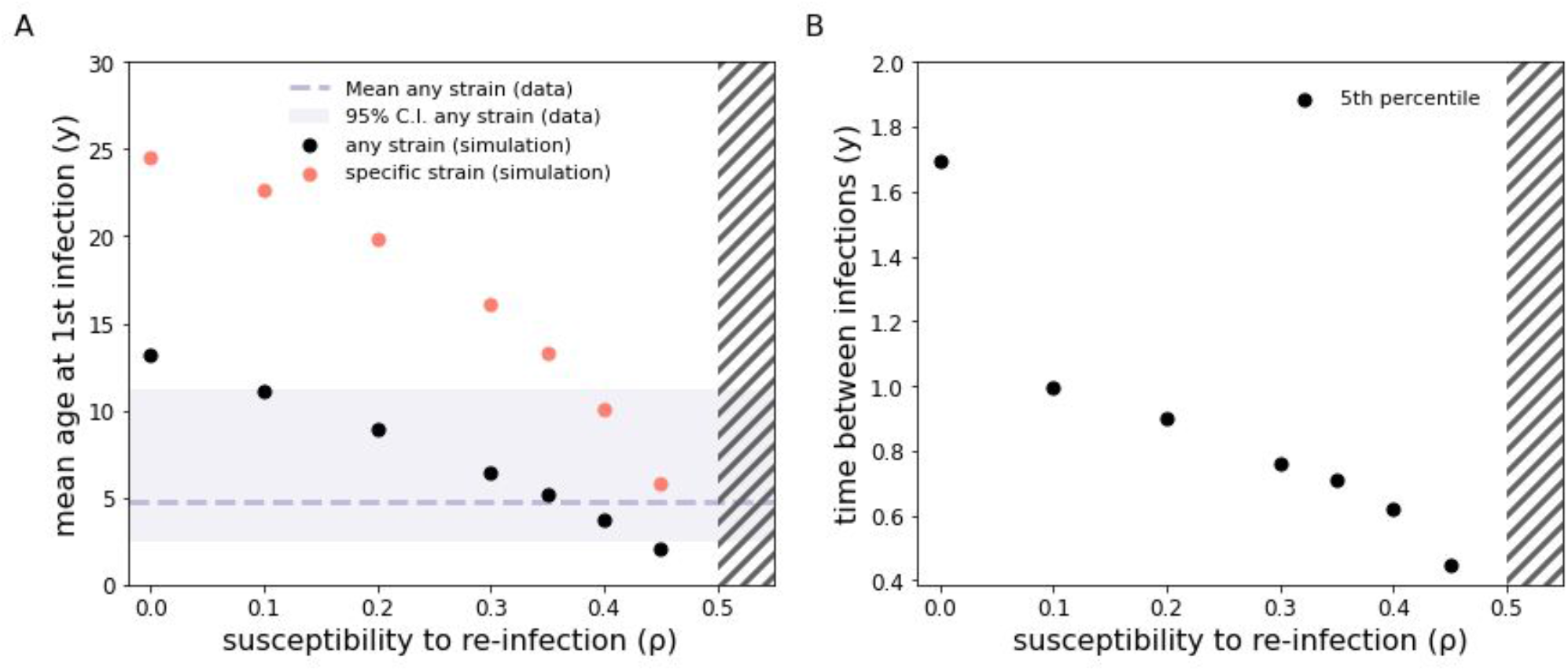
Impact of ρ on eHCoV dynamics and emergent strain hospitalisation rates. (A) Mean age at first infection with any eHCoV (black dots) and with a specific strain (red dots) as a function of ρ. The dashed line and band represent respectively the mean and 95% C.I. for the age at first infection with any eHCoV obtained from a pooled estimate (42). (B) 5th percentile of the time between consecutive infections by the same strain. Hatches indicate the ρ > 1/*R*_0_ region (for *R*_0_ = 2) where the dynamics are SIS-like (whereas for ρ < 1/*R*_0_ we observe epidemic behaviour). It should be noted that for ρ = 0 reinfection still occurs in our model because of external introductions, which we have assumed for simplicity to ignore pre-existing immunity to reinfection. Nonetheless, reinfection events induced by external introductions represent only a small fraction of all infection events. Epidemiological parameters are set to baseline values.

We investigated the role of susceptibility to reinfection (ρ) on the dynamics of eHCoVs. Increasing values of ρ (at fixed *R*_0_) yield a larger force of infection (FOI) which, in turn, affects almost every aspect of eHCoVs’ epidemiology. Figure 2A shows, for instance, the effect of ρ on the age of first infection with endemic strains: because of the relationship with FOI, larger values of ρ yield a younger mean age at first infection. Figure 2A also shows that values of ρ in the range [0.1 - 0.45] yield realistic values for the mean age at first infection with endemic strains, which is estimated to be 4.8 [2.5 - 11.2 95%C.I.] years globally (42). Consequently, in accordance with previous studies, our analysis rules out the possibility of complete, life-long complete immunity against reinfection by eHCoVs (i.e. ρ = 0) (42). Furthermore, very small values of ρ provide unrealistic age-specific incidence profiles. In the extreme case ρ = 0 (no reinfection), infections would occur only in children and young adults, contradicting empirical evidence of eHCoVs infecting older age classes (42).Finally, we note that values of ρ > 1/*R*_0_ cause a shift from an epidemic SIR-like behaviour to a stable SIS-like behaviour (43), suggesting values of ρ beyond ρ_*c*_ (indicated with hatches in Figure 2) are not epidemiologically plausible in the context of eHCoV dynamics.

Figure 2B also shows that our model reproduces reasonably short times between two consecutive infection events.Specifically, for ρ > 0.2 at least 5% of all reinfection events occur within 1 year since the last infection event. Our results agree with previous challenge experiments and cohort studies, which reported short-lived homotypic immunity (<1 year) against eHCoV reinfection in a minority of individuals (14,15,38).

### Modeling age-specific COVID-19 hospitalisation rates under eHCoV exposure dependence

We model individual-level heterogeneities in the probability of developing COVID-19 severe symptoms by assigning to each case a *severity score w*.

Exposure dependence is introduced by:

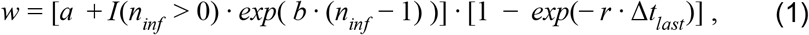

where *n*_*inf*_ is the number of previous infections to eHCoVs, Δ*t*_*last*_ is the time since the most recent first infection by any eHCoV and *I*(…) is an indicator function that equals 1 if the condition in the brackets is true and 0 otherwise. The two terms in square brackets reflect two distinct biological assumptions about the risk of developing COVID-19 severe symptoms following infection by the emergent strain:

1. Secondary infections with the same eHCoV do not provide any additional protection on the assumption that reinfection enhances homotypic immunity at the expense of cross-reactivity to SARS-CoV-2. After an individual encounters an eHCoV for the first time, the severity score is reset to 0, but increases progressively with time at rate *r* (waning of cross-protection) back up to the value *a* + *exp*(*b* · (*n*_*inf*_ − 1)).
2. The severity score increases exponentially with the number *n*_*inf*_ of previouinfections to any eHCoV to reflect the potential build-up of homotypic immunity superseding heterotypic immunity. The parameter *b* (boosting factor) quantifies the post-infection increment to the score, while *a* (baseline risk) represents the score in HCoV-naive individuals.

In each simulation, we sample a fraction n of infected cases without replacement, with severity scores representing sampling weights, and mark them as hospitalised (see Methods for additional details). For this theoretical exercise, we consider only individuals infected up to 50 days after the introduction of the emergent strain, i.e. those individuals that become infected during the early phase of the epidemic, before containment measures would have a significant impact on the epidemic. π is the overall fraction of cases hospitalised and thus represents the Infection Hospitalisation Ratio (IHR) which effectively aggregates multiple factors affecting reporting, e.g. visibility of symptoms, testing efforts, care-seeking behaviour and health care capacity.

Age-specific hospitalisation rates are shown in Figure 3 when varying *a* (baseline risk), *b* (boosting factor), *r* (waning of cross-protective immune responses) and π (IHR). Figure 3A-D shows that differences in the combination of these parameters can lead to widely divergent age patterns in hospitalisation rates. First, we note that the hospitalisation risk increases at older ages simply due to the dominance of strain-specific responses over cross-protective responses (i.e. *b* = 0, panels A,B). Second, for *a* > 0, the risk of hospitalisation is not a monotonic function of age. This is because when HCoV-naive individuals become infected with any eHCoV for the first time, their severity score for COVID-19 drops from *a* to 0, as illustrated by time trajectories of individual severity scores in Figure 3E; this explains why the risk of severe symptoms is so low in children and teenagers and also why it is slightly higher in the very youngest (0-5y) age class (44,45). Because the chances of encountering any eHCoV increase rapidly with age, most individuals have already encountered all eHCoVs by the time they reach adulthood. After that point, the average severity score increases at rate *r* (up to *a* + 1 in the absence of boosting), which results in increasing hospitalisation rates. Increasing values of *r* (waning of cross-protection) reduces this effect, since primed individuals revert to their maximal score faster. These observations hold also in the case where *b* > 0 (panels C,D); however, as expected, increasing values of the boosting parameter *b* result in a steeper increase in hospitalisation rates with age.

**Figure 3:**
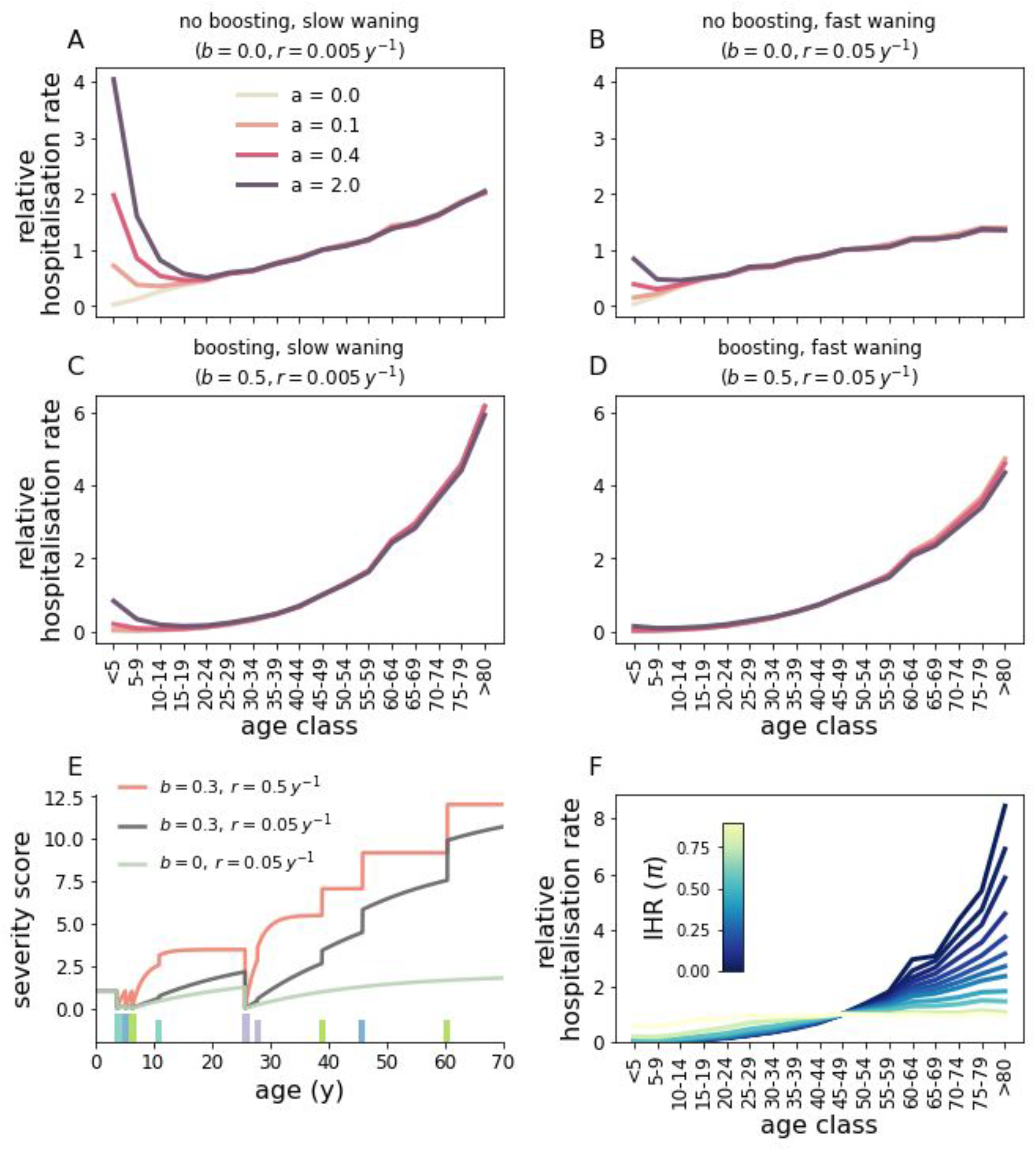
Impact of *b* (boosting factor), *r* (waning of cross-protection), *a* (baseline risk) and π (IHR) on age-specific hospitalisation rates. Panels (A-D) correspond to different combinations of parameters *b* and *r*. Within each panel, different curves correspond to different values of *a*. Here we assumed that a fraction π = 0.1 of all cases are hospitalised. For visualisation purposes, the rate corresponding to the 45-49y age range is set to one and the remaining rates are re-scaled accordingly. (E) Temporal evolution of the severity score for a single host under different combinations of *b, r* and *a* = 1. We considered three scenarios corresponding to no boosting and slow waning of cross-protection (green line, *b* = 0, *r* = 0.05 *y*), boosting and slow waning of cross-protection (black line, *b* = 0.3, *r* = 0.05 *y*), boosting and fast waning of cross-protection (red line, *b* = 0.3, *r* = 0.5 *y*). Bars indicate infection events, with each color corresponding to a different eHCoV. At birth, the score is identically equal to *a*. The score drops to 0 after encountering a new strain (thicker and taller bars), but increases thereafter at rate *r*. Secondary infections with the same eHCoV (smaller bars) do not provide any additional protection and only increase the score for *b* > 0. In panel (F) we set *a* = 0.4, *b* = 0.5, *r* = 0.05 *y*^−1^ and explore ρ. In all panels, epidemiological parameters are set to baseline values. Results are averaged over 50 samplings obtained from each of 5 different stochastic simulations (the impact of stochasticity on hospitalisation rates is further explored in Figure S2).

Figure 3F shows that, given a specific choice of *a, b* and *r* (and hence of a function for the severity score), smaller IHR (π) values lead to increasingly heterogeneous hospitalisation rates across age ranges. For very small values of the IHR, only those cases with the largest scores are hospitalised. In contrast, larger values of the IHR increase the number of infected cases that are hospitalised, which makes hospitalisation rates increasingly similar to age-specific attack rates. Crucially, this implies that the IHR has a non-linear impact on hospitalisation rates across ages and suggests that differences in IHR, perhaps due to heterogeneous capacity, admission policy and testing efforts, may partially explain inter-country variations in the relationship of age with hospitalisation (46).

In Figure 4A,B we compare model output and data for EU/EEA countries and the UK obtained from TESSy (ECDC source) (46) under our estimated epidemiological parameters (see Table 1) and a value of 5% for IHR, obtained after correcting counts of reported cases for non-uniform attack rates (47), similar to previously reported values (7,48,49).

**Figure 4:**
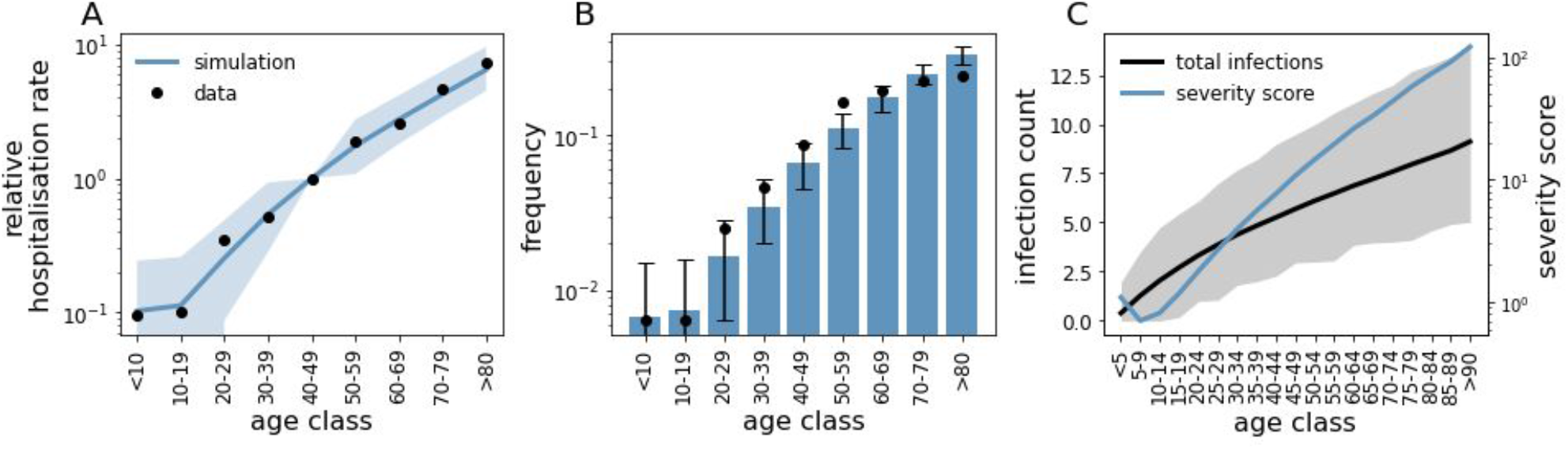
Comparison between model predictions and hospitalisation data for EU/EEA countries and the UK. (A) Simulated hospitalisation rates (blue line) and data (dots). The value corresponding to the 40-49y age range has been set to 1 for convenience and remaining values have been scaled accordingly. The shaded area indicates the 95% C.I. from simulations. (B) Age distribution of hospitalised cases in EU/EEA countries and the UK (dots) and mean distribution from simulations (bars). Error bars indicate the 95% C.I. from simulations. (C) Mean number of cumulative infections and severity score as a function of age (black and blue lines, respectively), at the time of the introduction of the emergent strain. Shaded area indicates the 95% C.I. from simulations. Here, we set ρ = 0.05, *a* = 1.5, *b* = 0.5, *r* = 0.05 *y*^−1^. Because our aim is mainly to illustrate the role of disease enhancement and cross-protection, we did not attempt to fit parameters *a* (baseline risk), *b* (boosting factor) and *r* (waning of cross-protection). Rather, we manually adjusted parameters in order to obtain a good visual agreement between data and simulations. Goodness of fit for chosen parameters was measured at *R*^2^ = 0.98. Results are averaged over 100 samplings from each of 50 different simulations. Other parameters are set to baseline values.

We obtain a good qualitative match to observed trends in hospitalisation rates and the age distribution of hospitalised cases (Figure 4A,B). In particular, the model seems to capture the relatively low rates observed in individuals aged 0-20 years and the rapid increase in hospitalisation rates after the age of 20. Age variations in severity scores (Figure 4C) underscore the important role of disease enhancement through repeated eHCoVs infections throughout life. Interestingly, Figure 4C also implies that children in the range 5-19y are less susceptible to severe symptoms than infants (<5 years), a pattern previously described for some countries such as Portugal, Italy, and the Netherlands (Figure S3) (44). As explained in the context of Figure 3, this optimum in protection from severe disease stems from the interplay between losing heterotypic responses in favour of homotypic responses with increasing exposure, and the protective effect of cross-protective immune responses after first exposure to an eHCoV. Figure S4 and Figure S5 further explore sensitivity of our results to susceptibility to reinfection and parameters defining the life-expectancy distribution, respectively.

We also explored the potential for sterilising (ie. infection blocking) heterotypic immunity to account for reduced risk of hospitalisation in children (8). However, levels of heterotypic immunity required to generate significant levels of protection were not compatible with the observed dynamics of eHCoVs (Figure S6).

### Modelling age-specific COVID-19 hospitalisation rates under age dependence

These data patterns observed for EU/EEA countries and the UK can also be recovered using a model in which disease severity depends only on an individual’s age (the age-dependent model). Briefly, this model assumes that the severity score is constant up to age *A*_0_, but increases exponentially with age thereafter:

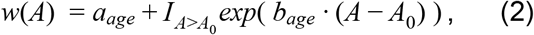

where *A* is age and *a*_*age*_, *b*_*age*_ are positive constants.

Figure 5A shows that such a model is able to capture the observed COVID-19 hospitalisation rates (*R*^2^ = 0.99), under the explicit assumption that individuals younger than *A*_0_ = 20 *y* are protected from severe symptoms. However, while both HCoV-exposure-dependent and age-dependent mechanisms seem to perform equally well in terms of age aggregated data, they yield different predictions about the individual risk of severe disease (Figure 5B). In particular, the former model predicts a small but non-negligible proportion of young individuals at high risk of severe COVID-19, whereas the age-dependent model does not. Figure 5C shows that heterogeneity in exposure to eHCoVs results in a fraction of young individuals displaying a severity score comparable to older individuals. In contrast, in the age-dependent scenario younger hosts display a systematically smaller and non-overlapping severity score compared to older ones (Figure 5D), requiring ad hoc assumptions on why severe disease can sporadically but still significantly happen in this age-group.

**Figure 5:**
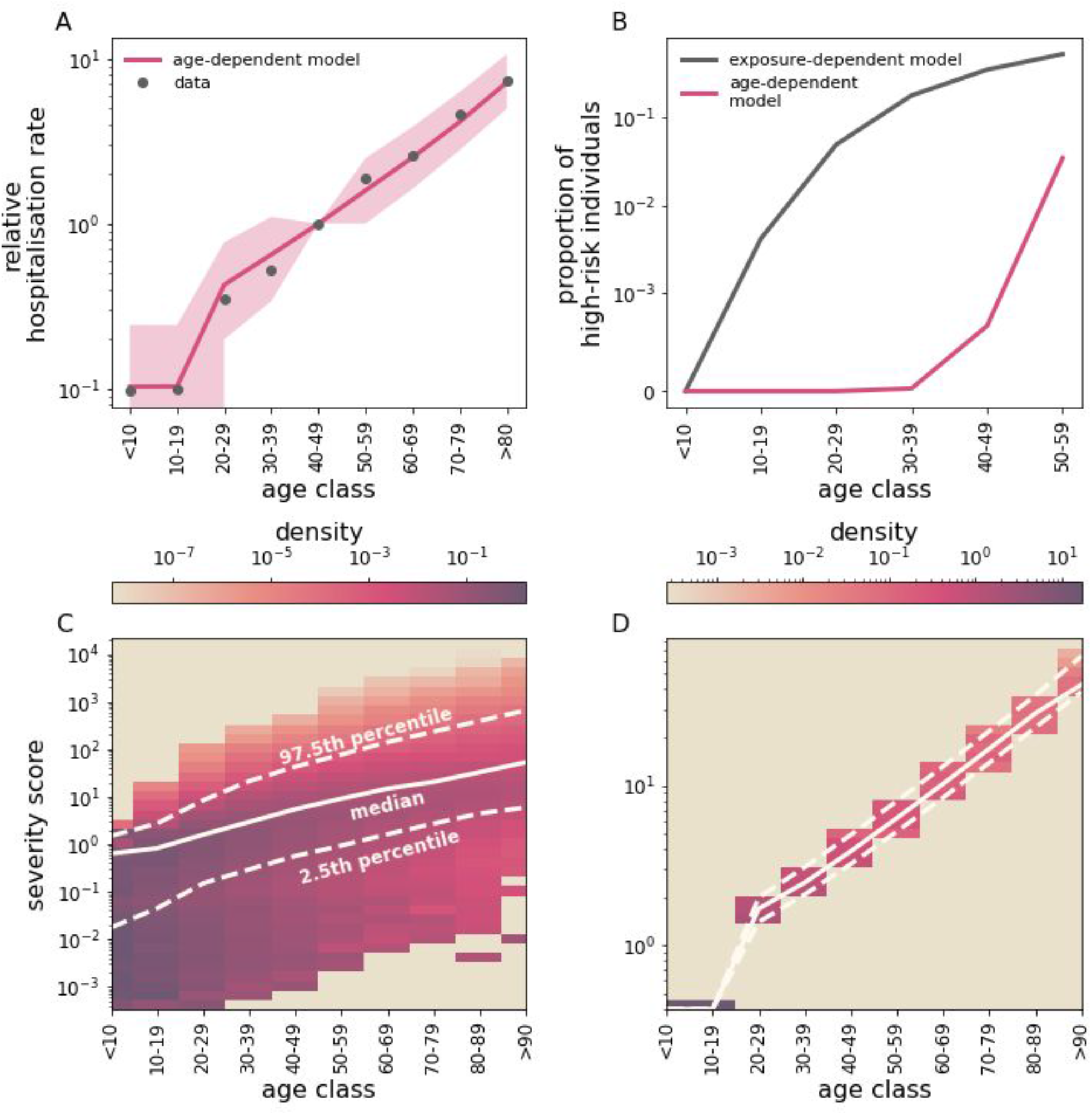
Implications of HCoV-exposure- and age-dependent-severity. (A) A model where disease severity depends only on age (the age-dependent model) is able to explain hospitalisation rates in EU/EEA countries andthe UK (dots). Line and filled area represent mean and 95% C.I., respectively. Here, we set *a*_*age*_ = 0.4, *b*_*age*_ = 0.052 *y*^−1^ and *A*_0_ = 20 *y*. Results are averaged over 100 samplings from each of 50 different simulations. (B) Individual risk of developing severe symptoms under exposure-dependent (black) and the age-dependent severity (fucsia). (C,D) Severity score distribution within each age class under exposure-dependant and age-dependent severity, respectively. Solid and dashed lines indicate the median score and the 95% percentile range, respectively. Please note that C,D have different scales. To estimate individual risk in (B), we first selected 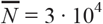 individuals completely at random in a single simulation (that is, we assume uniform infection rates across all age ranges) andthen sampled a fractionn = 0.05 of these 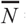 cases according to the sampling scheme outlined in the Methodssection. This operation was repeated 2 · 10^3^ times. Finally, we computed the proportion of high-risk individuals as the fraction of sampled cases whose risk is larger than the 25th percentile in the >60 years age range. In C,D, the score distribution is computed from a single simulation at the time of the introduction of the emergent strain. We set epidemiological parameters to their baseline values and *a* = 1.5, *b* = 0.5, *r* = 0.05 *y*^−1^.

## Discussion

Immunopathogenesis of COVID-19 is complex and still far from being completely understood (27). Here we offer a mechanistic explanation of age patterns of COVID-19 severity, based on individual exposure histories to eHCoVs. Our results support the notion that cross-protection induced by exposure to eHCoVs may explain the low frequency of COVID-19 severe symptoms in individuals under 20 years of age (8,45). If strengthening of homotypic immunity with repeated exposure interferes with the induction of cross-protective responses, this could explain why more immunologically experienced older age classes would be paradoxically more susceptible to COVID-19 disease upon first infection with SARS-CoV-2. Note that this increase of susceptibility with age due to reduced cross-protection is only a feature of a new epidemic; if SARS-CoV-2 becomes endemic, there should be sufficient homotypic immunity in older age classes to reduce the severity of a second infection.

Past exposure to eHCoVs may also act to exacerbate symptoms. Antibody Dependent Enhancement (ADE) (which is known to contribute to severity of secondary Dengue infections (50−53) and has been observed also in HIV (54,55), ebola (56,57) and influenza (58,59)) has been documented in SARS and MERS (60−64) but its role in the pathogenesis of COVID-19 is still unclear. Anti-spike IgG antibodies have been shown *in-vitro* to enhance ability to infect immune cells, notably macrophages, and induce the secretion of pro-inflammatory factors for both SARS-CoV (65) and SARS-CoV-2 (66). Recent studies also indicate widespread T cell reactivity in blood samples obtained during the pre-pandemic period (20,21,23). Pre-trained T cell immunity is likely generated by previous exposure to eHCoVs (67,68) and is generally thought to promote viral clearance (20,69); however, dysregulated CD4 T cell responses have also been shown to contribute to cytokine storm in severe COVID-19 patients (70).

The age distribution of COVID-19 may thus be explained either by the decay of protective cross-reactive responses to eHCoVs or by the accrual of non-protective cross-reactive responses. However, these are unlikely to be the sole drivers of COVID-19 severity. Indeed, we find that a simple age-dependent model can also match empirical hospitalisation rates in the other age-groups (Figure 5) but only under the explicit assumption that individuals below 20 years are intrinsically protected against severe illness, for example due to age differences in ACE2 expression in the respiratory tract (71). In contrast, the exposure-dependence model shows that protection against severe symptoms in children and teenagers can emerge dynamically due to the existing, age-dependent immunity landscape present in the population before emergence of SARS-CoV-2. This scenario permits a small but non-negligible proportion of the young to behave as outliers in terms of their immunity status and are indeed expected to be at increased risk of severe disease at levels comparable to that of older individuals (Figure 5), and may explain why certain children presenting no underlying health conditions have been reported to develop severe symptoms associated with SARS-CoV-2 infection (45,72,73). Although data is limited, the age distribution of severe eHCoVs infections contrasts with the age distribution of severe COVID-19, following a pattern of reduction in severity through repeated exposure (Figure S7). We note that this supports the hypothesis that frequent exposure to eHCoVs throughout age may favour homotypic immune responses to those viruses, thereby compromising the development and persistence of cross-reactive responses that could reduce the severity of disease with the newly emerging coronavirus.

In this work, we considered reported hospitalisation rates as a proxy for severity of symptoms. Alternatively, we could have considered other indicators of disease severity, e.g. rates of severe hospitalisations and fatalities. The former, however, is particularly sensitive to local capacity. The apparent decline in ICU admissions observed in older age-groups, for example, is likely driven by clinical decisions and capacity, rather than by a true decline in disease severity (74). Fatality rates, on the other hand, are unlikely to provide a robust signal at younger ages because of the small numbers of lethal outcomes in children and teenagers (72). We focused on data aggregated at the European level, noting that individual countries show a qualitatively similar behaviour (Figure S3). In principle, inter-country variation in age-specific hospitalisation rates might stem from differences in eHCoV circulation patterns. However, in sensitivity exercises, we have shown that heterogeneities in testing and containment efforts, as measured by the IHR (i.e. π in our framework), can affect the shape of hospitalisation rates, even under the same biological and epidemiological conditions (Figure S8). Disaggregating these factors from biological mechanisms of pathogenesis will be essential in further research to better understand the human immune responses against SARS-CoV-2 in the context of immunological cross-reactions induced by previous exposure to eHCoVs.

## Methods

### Additional details on multi-strain model

We consider a homogeneously mixed host population of constant size N. The population is endowed with a realistic age profile modelled using a Weibull distribution with scale θ_*a*_ and shape *k*_*a*_ (75).

We consider a multi-strain epidemic model with *n* strains. Each strain *i* is characterized by aper-contact transmission probability β_*i*_ (*i* = 1, 2, …, *n*) and a daily recovery probability σ_*i*_. Asreinfection is commonly observed in eHCoVs (15,37,38), we assume that the recovered status from strain i provides only partial protection against reinfection with the same strain; in particular, we assume that exposure to a previously encountered strain results in infection with probability ρ. The case ρ = 0 corresponds to complete, life-long immunity upon recovery. For simplicity, weassume that strains do not interact with each other and therefore spread independently. Infection and recovery processes are stochastic and occur in discrete time, with the time unit set to 1 day.

Because eHCoVs display marked annual incidence patterns (76,77), we add an external sinusoidal forcing *f*(*t*) to transmissibility with period 1 year and intensity ε :

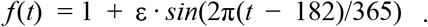

If we assume that each individual establishes on average *k* daily contacts, the overall force of infection associated to strain *i* is given by:

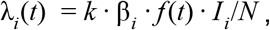

where *I*_*i*_ is the number of individuals infected with strain *i*.

In this work we consider *n* = 5 strains; strains labelled *i* = 1, 2, 3, 4 represent eHCoVs, while strain *i* = 5 represents the emergent SARS-CoV-2. eHCoVs are introduced into the system at *t* = 0, while the emergent strain is introduced at a later time *T*_*inv*_ by infecting 10 individuals chosen at random. We choose *T*_*inv*_ to be large enough (here we set *T*_*inv*_ = 160 *y*) so that by the time the emergent strain is introduced, both population demography and eHCoVs have already reached stationarity. We avoid permanent extinction of eHCoVs by allowing external introductions, which occur at an individual rate v.

### Modeling hospitalisation with heterogeneous risk of severe disease

Let *X* = {*x*_1_, *x*_2_, …, *x*_*n*_} be a list of hosts infected by SARS-CoV-2 over a particular time window in a single simulation. In order to select hospitalised cases from *X*, we first draw the total number *m* of hospitalised individuals, which is binomially distributed with parameters *n* and π. In a second step, we create a list 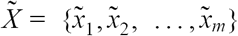 of hospitalised cases by randomly selecting *m* cases without replacement from *X*, with odds proportional to their corresponding scores *w*_1_, *w*_2_, …, *w*_*n*_.

## Data Availability

Data from The European Surveillance System - TESSy, provided by Austria, Croatia, Cyprus, Estonia, Finland, Germany, Hungary, Ireland, Italy, Latvia, Lithuania, Luxembourg, Malta, Netherlands, Norway, Poland, Portugal, Slovakia, United Kingdom and released by ECDC.

https://covid19-surveillance-report.ecdc.europa.eu/

## Acknowledgements

FP was funded by the UKRI GCRF One Health Poultry Hub (Grant No. BB/S011269/1), one of twelve interdisciplinary research hubs funded under the UK government’s Grand Challenge Research Fund Interdisciplinary Research Hub initiative. JL was supported by a Lectureship from the Department of Zoology, University of Oxford. SG acknowledges funding from the ERC ‘UNIFLUVAC’ (812816) and MRC CiC 6 as well the Georg und Emily Von Opel Foundation. DSCD is funded by the grant 19/23343-7 and 2020/06160-3 from the Säo Paulo Research Foundation (FAPESP). MG is supported by Fundaçäo de Amparo à Pesquisa do Estado do Rio de Janeiro (FAPERJ). LCJA acknowledges funding from PAHO (Pan American Health Organization) SCON2019-00572 (SCON2018-00572). UO and PW declare no funding. FP and JL thank Dr. Craig Thompson and Prof. Paul Klenerman for useful discussion.

## Authors’ contributions

SG, UO and JL conceived the idea. FP and JL developed the model. FP implemented the model and ran simulations. FP, JL analysed model output and conceived the visualisation of results. FP, SG and JL led the writing, with contributions of UO and PW. All authors contributed to text revisions and accepted the submitted version.

## Disclaimer

The views and opinions of the authors expressed herein do not necessarily state or reflect those of ECDC. The accuracy of the authors’ statistical analysis and the findings they report are not the responsibility of ECDC. ECDC is not responsible for conclusions or opinions drawn from the data provided. ECDC is not responsible for the correctness of the data and for data management, data merging and data collation after provision of the data. ECDC shall not be held liable for improper or incorrect use of the data.

